# Crop Filling: a pipeline for repairing memory clinic MRI corrupted by partial brain coverage

**DOI:** 10.1101/2023.03.06.23286839

**Authors:** Gonzalo Castro Leal, Tim Whitfield, Janaki Praharaju, Zuzana Walker, Neil P. Oxtoby

## Abstract

Data-driven solutions offer great promise for improving healthcare. However standard clinical neuroimaging data is subject to real-world imaging artefacts that can render the data unusable for computational research. T1 weighted structural MRI is used in research to obtain volumetric measurements from cortical and subcortical brain regions. However, clinical radiologists often prioritise T2 weighted or FLAIR scans for visual assessment. As such, T1 weighted scans are often acquired but may not be a priority. This can result in artefacts such as partial brain coverage being systematically present in memory clinic data.

Here we present a neuroimaging pipeline to ameliorate such situations by filling the missing regions with synthetic data. We validate on artificially cropped scans from the Alzheimer’s Disease Neuroimaging Initiative (ADNI), showing that our pipeline largely removes the artefact, improving volumetric biomarker accuracy while also retaining statistical differences between diagnostic groups. We demonstrate utility by achieving diagnostic classification performance comparable to uncorrupted data. This is an important contribution towards moving research from the lab into the real world.

## Introduction

Alzheimer’s disease (AD) is a neurodegenerative condition that affects over 9 million people in Europe and has a prevalence of 60-80% among the population living with dementia around the world [1]. The contemporary approach to characterising AD in research settings comprises the assessment of aggregated β-amyloid (A) and pathological tau (T), as well as neurodegeneration (N). Non-invasive imaging techniques used to assess these have been described under the AT(N) research framework proposed by the NIA-AA in 2018 [2]. According to this framework, neurodegeneration is best measured by structural magnetic resonance imaging (sMRI), as volumetric measurements of brain regions of interest can be accurately obtained from T1 weighted (T1w) sequences.

The development and improvement of semi-automated tools for brain segmentation have allowed for quantitative analysis of neurodegeneration using MRI scans from large cohorts in a significantly shorter time compared to manual segmentation. This has also enabled the analysis of multiple and/or more specific regions of the brain. Some of the most popular software implementations include, but are not limited to, FreeSurfer (Martinos Center for Biomedical Imaging, Harvard-MIT, Boston; https://surfer.nmr.mgh.harvard.edu/) and FSL (Analysis Group, FMRIB, Oxford, UK; https://fsl.fmrib.ox.ac.uk/fsl/fslwiki). One of the features shared between such brain segmentation tools is the use of whole brain atlases or templates. For example, FreeSurfer uses brain topology to establish the boundaries of cortical regions [3], while FSL-FIRST relies on a whole-head registration to the MNI152 space [4]. These tools have been used extensively to study AD from different perspectives: for disease clustering and prognosis [5-7], to establish the diagnostic utility of different brain regions [8-11], and to estimate regional brain atrophy trends/rates during disease progression [12, 13].

Image data used in semi-automated brain segmentation tools should be carefully selected, as different factors can affect the accuracy and reliability of the results. Quality controls often include assessment of technical artifacts such as head coverage, radiofrequency noise, signal inhomogeneity, and susceptibility, as well as motion artifacts like blurring and ringing [14]. Such technical artifacts are more common in real-world data than in the highly controlled clinical research studies typically leveraged by researchers to develop new quantitative methods for neuroimage analysis. Among these factors, partial brain coverage was by far the most prevalent one encountered in the CODEC (https://ucl-codec.github.io) dataset of routinely collected data from the Essex Memory Clinic, near London in the United Kingdom [15]. This was due to scans from the memory clinic prioritising visual ratings on T2-weighted MRI, with the T1w scans field of view (FOV) reduced due to time efficiency considerations.

Here we report on our investigation into cropping-induced artefacts in T1w sMRI volumetric measurements obtained from FreeSurfer and our proposed and validated solution that enables real-world memory clinic data to be used for quantitative research and development. The paper is structured as follows. The following section describes the data we analysed, our new crop-filling pipeline, and our experimental design including the statistical methodology used to assess the pipeline. In the results section we discuss our findings, focusing on the most affected regions in the early stages of AD, and a group analysis across all affected regions. In the discussion we summarise the key results and present limitations and future work. The final section closes with a conclusion and outlook.

## Materials and Methods

Participants were selected from the Alzheimer’s Disease Neuroimaging Initiative (ADNI; https://adni.loni.usc.edu/) observational research study, and the CODEC dataset was used to inform the cropping artefact and the clinical scenario. The ADNI sample included all participants having both a T1w and T2w 3D 1.5T MRI scan at the baseline visit, which included 173 cognitively normal (CN); 262 with Mild Cognitive Impairment (MCI); and 114 diagnosed with probable AD dementia. The T1w and T2w MRI scans had resolutions of 1×1×1.25 mm and 1×1×3 mm, respectively. The CODEC sample comprises real-world neuroimaging and clinical data, including neurocognitive test scores. The 3D MRI acquisitions are T1w (1×1×1 mm), T2w (0.5×0.5×5 mm) and FLAIR (0.5×0.5×5 to 0.9×0.9×6 mm) scans. The FOV of T1w sequences is 120×256×224 mm. The ADNI data were pre-processed to resemble the CODEC memory clinic data, which included artificially cropping the T1w scans to a lateral FOV of 120 mm and down-sampling the T2w scans to have an axial slice thickness of 5 mm. Some demographics of included ADNI participants are summarized in Table 1.

**Table 1:** Participant demographics from ADNI.

|  | CN | MCI | AD |
| --- | --- | --- | --- |
| N (Female) | 180 (89) | 274 (108) | 123 (58) |
| Age (Mean ± STD) | 75.34 ± 4.87 | 74.19 ± 7.07 | 74.43 ± 7.74 |
| MMSE (Mean ± STD) | 29.1 ± 1.0 | 27 ± 1.80 | 23.13 ± 2.10 |

### Experiments

The proposed pipeline flow-chart is displayed in **Figure 1**. It fills a cropped T1w MRI with voxels synthesized from a T2w sMRI using FreeSurfer’s ‘*SynthSR*’ and ‘*SynthSR Hyperfine*’ tools [16, 17]. Both tools output synthetic T1w MPRAGE 1mm isotropic scans from a given input of any contrast or spatial resolution. The former could use a T2w or FLAIR scan while the later uses the combination of a T1w plus a T2w to give a more refined output. In the process of filling the missing data, registration and resampling are often needed. The “Registration” steps use FreeSurfer’s ‘*mri_robust_registration*’ [18]. “Filling” involves resampling synthetic images to the cropped scan spatial resolution and adjusting shape, prior to filling the edges of the image with the recovered data. The resampling was performed using FreeSurfer’s ‘*mri_convert*’ tool.

**Figure 1:**
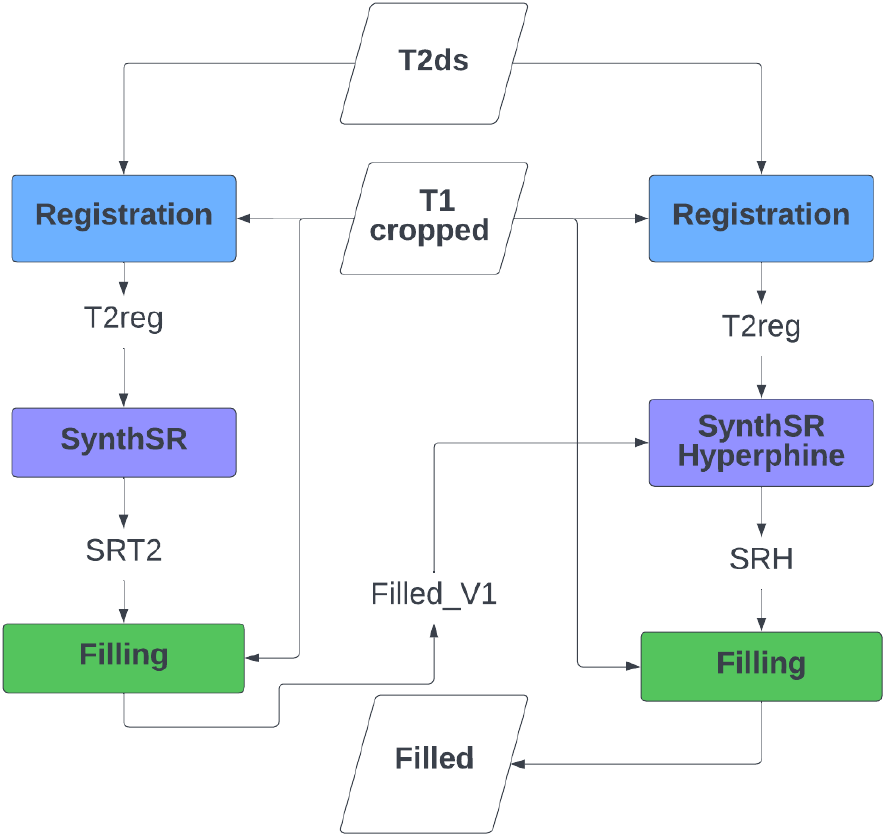
Filling pipeline, depicting every step. “T2ds” is the down-sampled T2w scan, “T1 cropped” is the artificially cropped T1w scan, “SRT2” is the synthetic image obtained from ‘*SynthSR’*, “SRT2reg” is the SRT2 image registered to the T1 cropped, “Filled_V1” is the first filled image obtained by filling the missing data in the T1 cropped scan with the SRT2reg image, “SRH” is the synthetic image obtained from ‘Hyperfine SynthSR’, “Filled” is the final result from the pipeline.

All images were then processed with ‘*recon-all*’ [19-27] from FreeSurfer version 7.1.1 (https://surfer.nmr.mgh.harvard.edu/fswiki/rel7downloads). This pipeline involves motion, intensity and bias field correction, skull stripping, volumetric labelling and registration, grey/white matter segmentation and registration to predefined atlases. All scans considered for this study had passed a quality check performed by ADNI when analysed with previous versions of FreeSurfer, so no further visual inspection for segmentation errors was performed. The volumetric measurements of the cortical regions, defined by the Desikan-Killiany atlas [3] stored in the aparc.stats file, and subcortical structures, from the aseg.stats file, were compared. To reduce dimensionality in the analysis, cortical segmentations were grouped into their respective cortical lobes according to the Klein and Tourville [28] description.

### Statistical Analysis

Correlation, bias and statistical tests were used to evaluate regional brain volumes obtained from cropped, crop-filled and hyperfine synthetic (SRH) MRI volumes against the ground truth volumes from unprocessed (the original ADNI images). Correlation was assessed as *r*^*2*^ where *r* is the Pearson’s coefficient. Bias is reported as mean percentage error. Pipeline performance was also assessed using statistical tests for group differences both within and between diagnostic groups — to ensure that disease signal was not affected by the artefact removal process.

Student’s t-test was used for intra-group comparisons (original vs processed within CN/MCI/AD respectively). Welch’s t-test was used for the comparison of original CN vs processed MCI/AD groups. We controlled for age, sex, and head size (intracranial volume) in the statistical tests using robust linear models.

### Classification Analysis

We also investigated the influence of cropping (and filling) on an example clinical application of interest: classification of diagnostic groups using regional brain volumes in Support Vector Machines (SVM) [29]. ANOVA was used to find the best combination of features for each classification task: CN vs AD, CN vs MCI and MCI vs AD. The best combination of parameters (kernel: linear, polynomial, or radial basis function [RBF]; gamma, C, degree, and nu) was evaluated through Bayesian Optimization. Repeated 10-fold cross-validation was used to evaluate classification performance with training performed on ground truth data and testing on cropped, filled and ground truth data. Both AUC and balanced accuracy are used as metrics for performance comparison.

## Results

**Figure 3** shows cropping severity for regions that lost on average more than 3% of volume. The reduction in the FOV affected the left lobe (orange box plots) slightly more than the right (blue) in the middle temporal region for all diagnostic groups and the superior temporal region for CN and MCI groups (all p values < 0.05, Student’s t-test).

**Figure 2:**
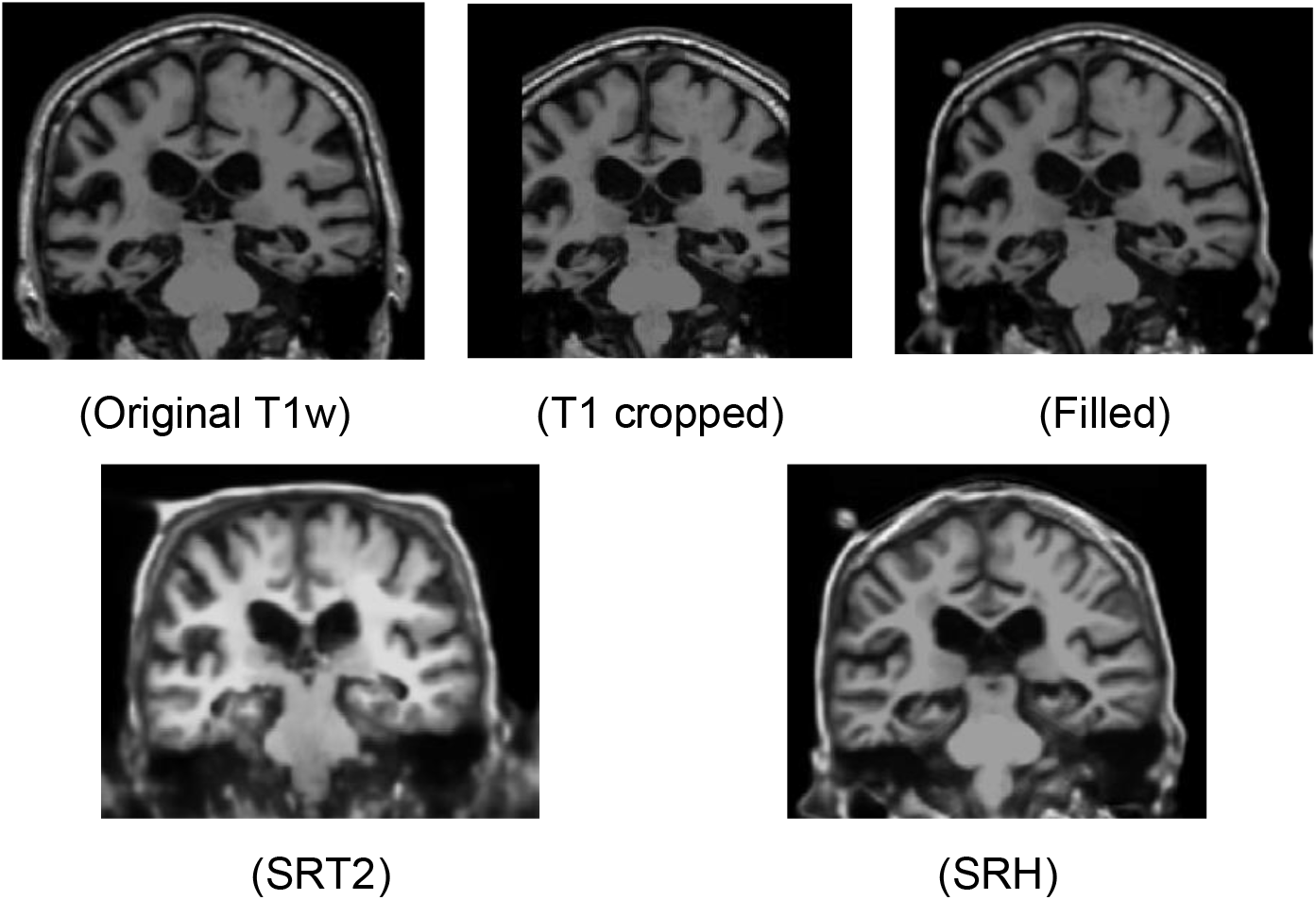
The original (Original T1w), artificially cropped (T1 cropped) and filled (Filled) scans in the top row. The synthetic T1w image (SRT2) obtained from only the T2w scan with ‘SynthSR’, and the hyperfine synthetic one (SRH) from ‘Hyperfine SR’ in the bottom row.

**Figure 3:**
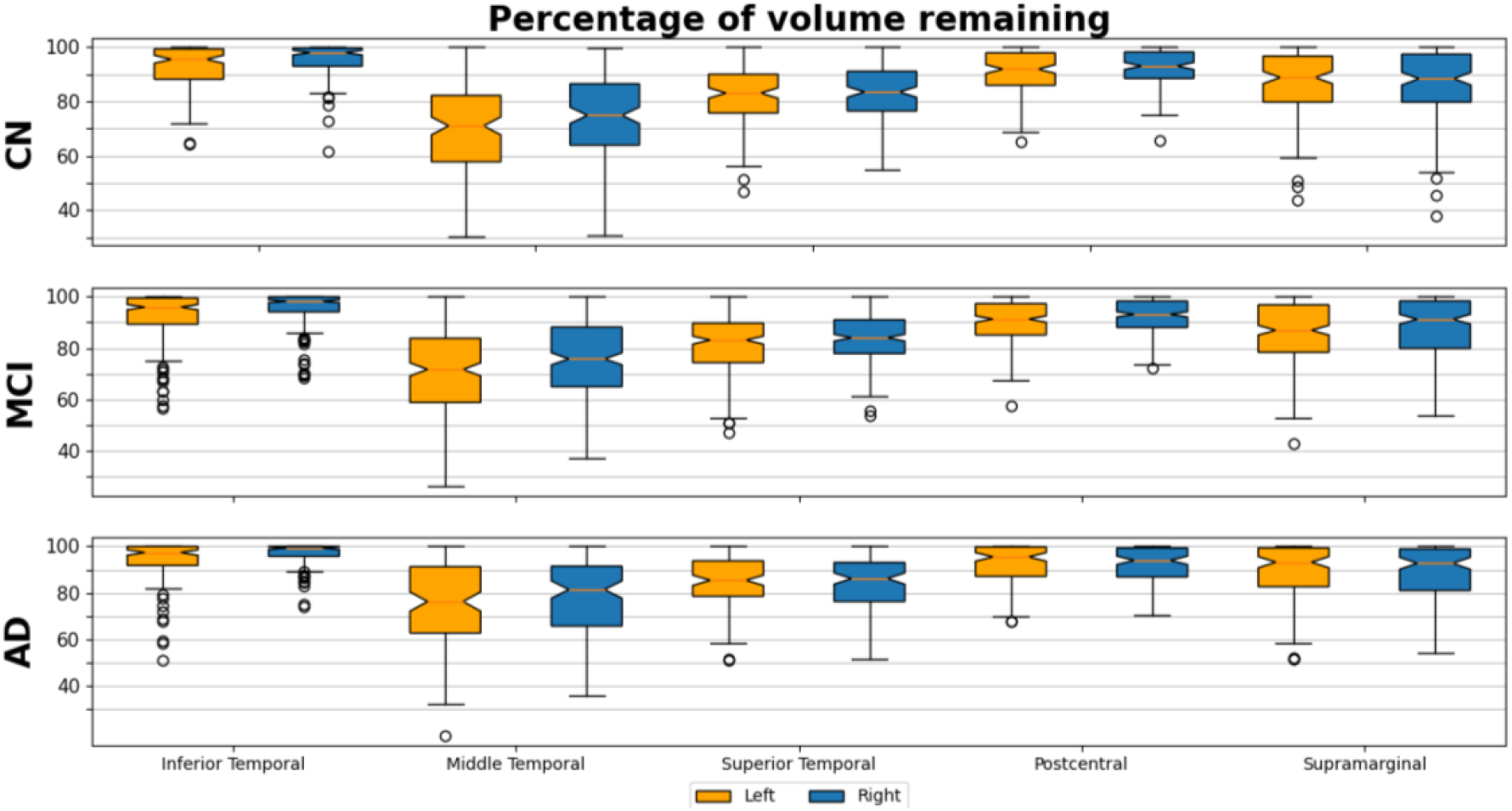
Cropping severity in affected regions. Box plots of the remaining percentage of volume in cropped T1w sMRI across diagnostic groups. Left hemisphere (orange) and right hemisphere (blue).

The results of correlation and bias within diagnostic groups are summarized in **Figure 4** for cropped, filled and synthetic scans. Only three AD-specific regions of interest are displayed (ventricles, temporal lobes, and hippocampus), in addition to the combined metrics from cortical and subcortical structures. Fully synthetic images had the worst results overall, except in the temporal lobes (cropped images were worst), with filled images consistently showing high correlation and low bias.

**Figure 4:**
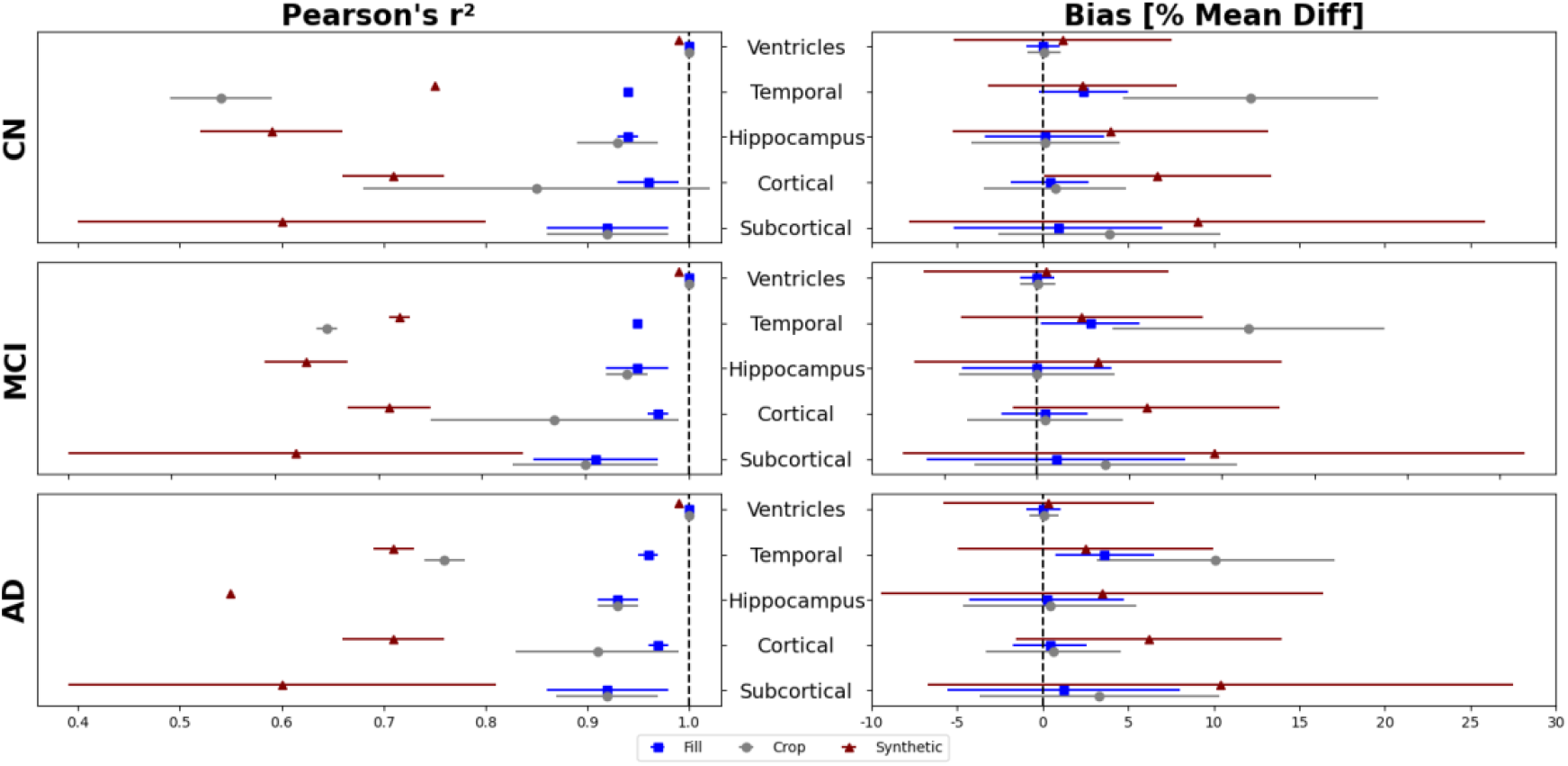
Bias and correlation plotted as value ± standard deviation.

The statistical differences between original and processed scans are depicted visually in **Figure 5** and **Figure 6. Figure 5** shows that crop-filling (right) almost completely recovers ground truth volumes affected by cropping (left), in each diagnostic group. **Figure 6** shows that regional disease signal (CN vs AD and CN vs MCI) corrupted due to cropping (1^st^ and 3^rd^ rows) is almost completely recovered by our crop-filling pipeline (2^nd^ and 4^th^ rows).

**Figure 5:**
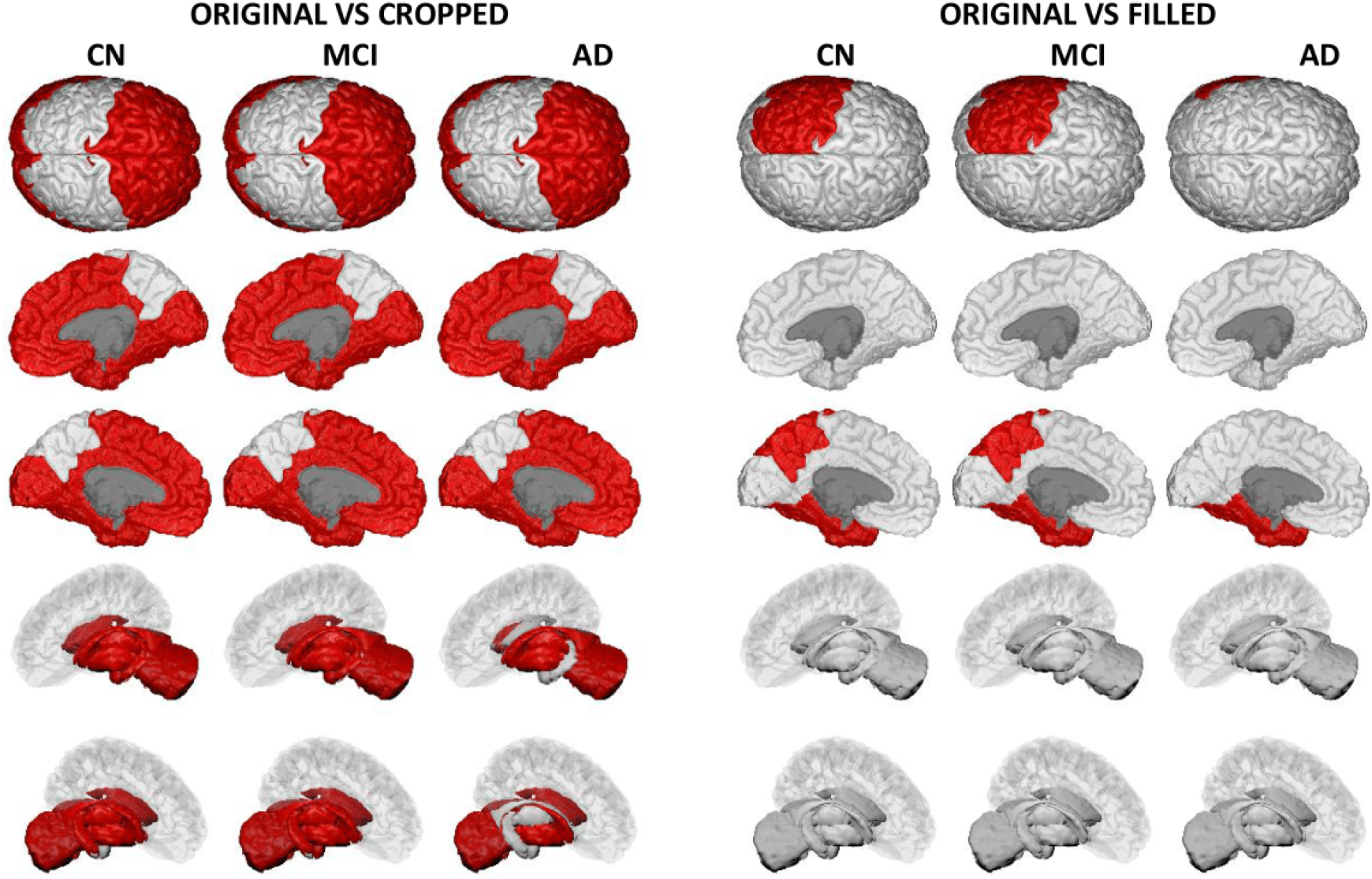
Brain-painter [30] outputs displaying the regions with a *p-value* < 0.05 in red as a result of the t-test performed between original data and cropped/filled data. Looking at the last four rows, from top to bottom, the odd ones include the right hemisphere and, the even ones the left.

**Figure 6:**
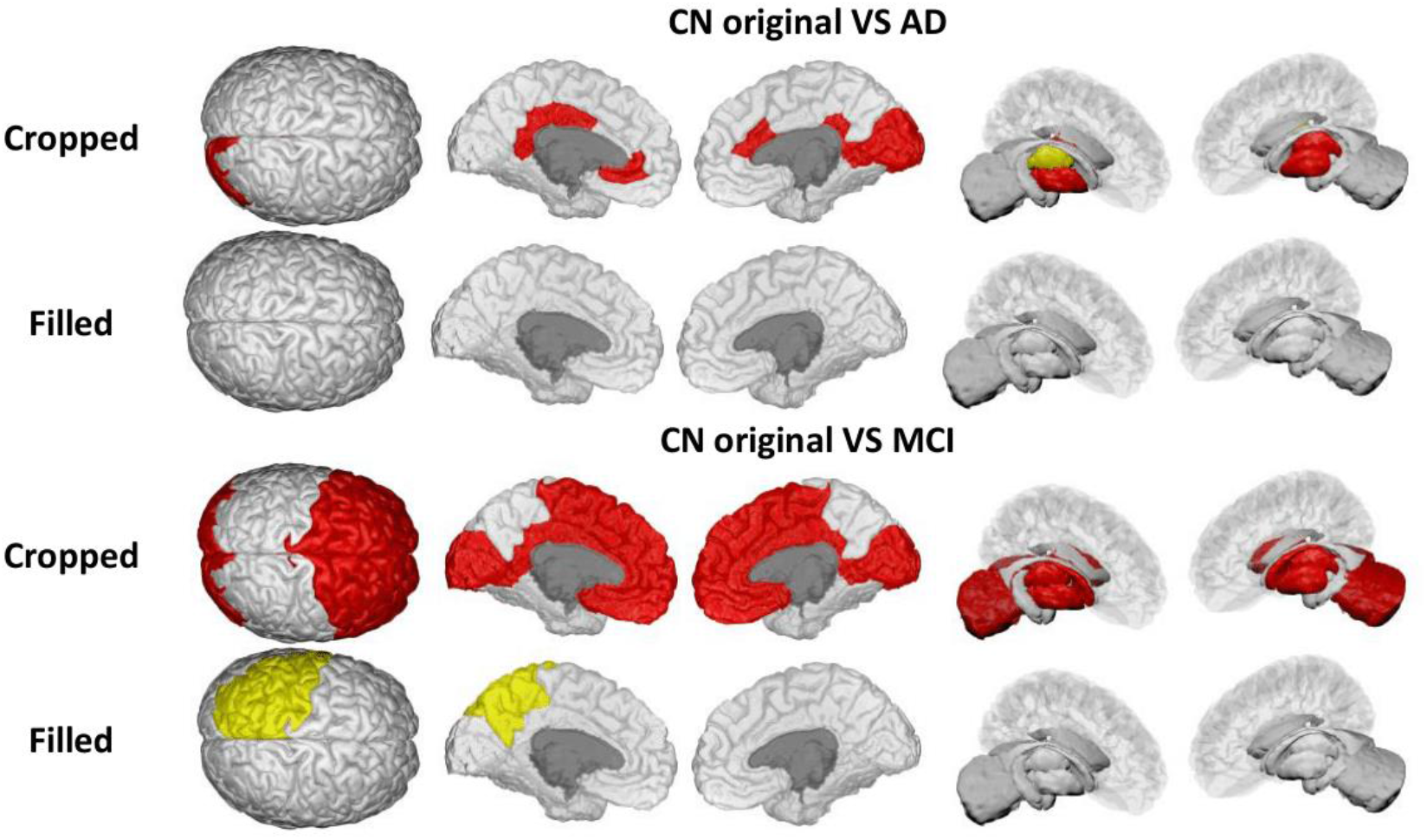
Brain-painter [30] outputs highlighting the loss (gain) of significant difference in regions of the brain that are (are not) supposed to be significantly different according to t-test performed between CN and AD/MCI original data. Red indicates a severe change, while yellow indicates a milder alteration. There is some asymmetry.

**Table 2** shows AUC and balanced accuracy for each classification task (CN vs AD, CN vs MCI and MCI vs AD) using original, cropped, and filled data. Cropping reduced clinical classification performance to the level of random guessing, and our crop filling pipeline almost completely repairs this. Feature selection (see Methods) chose a unique combination of regional volumes for each classification task. CN and MCI were best separated by using volumetric measurements of left entorhinal cortex, bilateral hippocampus, and bilateral amygdala. CN and AD subjects were best separated by using left inferotemporal, left entorhinal, right middle temporal cortexes, bilateral hippocampus, and bilateral amygdala. MCI and AD subjects are best separated by using bilateral inferotemporal, right fusiform gyrus, right hippocampus, and right amygdala. These findings align well with what is described about the progression of AD according to Braak’s stages [31], with earlier stages (CN vs MCI) involving the entorhinal cortex and middle to late stages (MCI vs AD) involving the fusiform gyrus and other temporal regions (inferior, middle and superior).

**Table 2:** AUC and Balanced Accuracy for each of the classification tasks on the different volumetric data.

|  | Data | AUC | Balanced Acc. |
| --- | --- | --- | --- |
| CN vs MCI | Original | $0.80 \pm 0.06$ | $0.71 \pm 0.06$ |
| | Filled | $0.78 \pm 0.06$ | $0.70 \pm 0.06$ |
| | Cropped | $0.54 \pm 0.04$ | $0.52 \pm 0.08$ |
| CN vs AD | Original | $0.93 \pm 0.04$ | $0.88 \pm 0.06$ |
| | Filled | $0.93 \pm 0.05$ | $0.87 \pm 0.07$ |
| | Cropped | $0.55 \pm 0.10$ | $0.54 \pm 0.09$ |
| MCI vs AD | Original | $0.67 \pm 0.10$ | $0.62 \pm 0.07$ |
| | Filled | $0.67 \pm 0.11$ | $0.60 \pm 0.06$ |
| | Cropped | $0.50 \pm 0.10$ | $0.50 \pm 0.06$ |

## Discussion and Limitations

This work focussed on developing a pipeline that can solve the systematic reduction in the FOV of T1w MRI scans (“cropping”) seen in real-world data from a memory clinic in the UK. This was motivated by allowing such data to be used in quantitative research and development, which is critical for addressing the current lack of ethnic and sociodemographic diversity [32, 33] in the large neuroimaging data sets such as ADNI that are currently driving innovations including data-driven disease progression modelling [34, 35]. The cropping issue is essentially a missing data problem, and thus the approach implemented was to recover the data in a sensible way.

Our experiments focussed on recovering image-derived features in a routinely collected imaging modality relevant to Alzheimer’s disease progression modelling, i.e., regional brain volumes from T1w MRI. To validate the proposed pipeline, we used ADNI data as ground truth and artificially cropped T1w sMRI and down-sampled T2w MRI to mimic the real-world data. The method made use of other FreeSurfer tools, ‘*mri_robust_registration*’ and ‘*SynthSR*’, to estimate the missing data from both scan modalities and fill-in the gaps to obtain a “filled” version of the original scan. Original, cropped and filled scans were then processed with FreeSurfer ‘*recon-all*’ tool, and the results were analysed.

The loss of FOV mostly affected a few cortical regions from the temporal and parietal lobes, with the middle temporal cropped by an average of 24% and 28% in the right and left hemispheres, respectively. In general, the left hemisphere was significantly more cropped than the right. This could be specific to the ADNI scanning protocol and deserves further investigation. As might be expected in a neurodegenerative disease, the percentage of tissue preserved increased with disease severity from CN to MCI to AD groups.

Remarkably, FreeSurfer-estimated volumes of subcortical structures were also affected by the cropping of the cortical regions. This might be due to FreeSurfer’s use of the whole brain when parcellating all regions using recon-all. The *average* bias and correlation for estimated subcortical volumes were comparable between cropped and filled scans, but the *variance* in bias was significantly higher in cropped scans, showing that crop-filling can reduce this source of variation that would likely confound quantitative statistical analyses of disease progression.

Our results show that the crop filling pipeline almost completely recovers artifact-corrupted disease signal. For example, cropping removes group differences in regional brain volumes between diagnostic groups (e.g., frontal lobe grey matter volume CN vs MCI uncropped), or introduces statistically significant group differences where they are known not to exist (e.g., cingulate grey matter volume MCI vs AD uncropped).

For practical demonstration of how crop-filling can help with statistical analyses of disease progression in real world data, we performed classification experiments using support vector machines. Our results showed that data from cropped scans severely reduced classification performance to little better than random guessing of diagnosis, whereas crop-filled data matches ground truth data performance (in terms of AUC and balanced accuracy).

Our study has some limitations that open avenues for important future work in training disease progression models for real-world applications in dementia. Firstly, there might exist other tools for performing steps in our pipeline that could improve results, including emerging deep-learning methods for robust registration (MONAI – https://monai.io/) and replacing FreeSurfer’s ‘*SynthSR*’ tool with an alternative image synthesiser such as DANI-Net [36]. Secondly, different models/methods of assessing disease progression might require alternative performance metrics — for example, structural similarity metrics might be more appropriate for voxel-wise analyses. Third, the imperfect matching of ADNI MRI protocols to the real-world data might confound our results (although we don’t think so), so future work will explore using real-world data in otherwise unchanged experiments (artificial cropping, etc.) once the reduced FOV has been corrected in the hospital protocol. This would also address the possibility that the ADNI research data may have a higher SNR than real-world data. Fourth, image synthesis may benefit from adding other imaging modalities such as FLAIR.

## Conclusion

Motivated by artefacts seen in real-world data, we have presented a new method to recover missing voxels from brain MRI having partial coverage (lateral cropping) due to a reduced field of view. Our pipeline leverages freely available tools in the FreeSurfer software suite and was validated in publicly available data from the ADNI study that was artificially cropped to match the observed real-world artefacts. Experimental results show that the pipeline recovers the missing data almost completely, and in a manner useful for disease progression modelling. Making real world neuroimaging data useable for research and development in this manner is an important step towards the goal of improving inclusivity and diversity in computational medicine and healthcare.

## Data Availability

ADNI data is available from adni.loni.usc.edu

https://adni.loni.usc.edu

## Acknowledgements

This research was supported by the National Institute for Health Research University College London Hospitals Biomedical Research Centre. NPO is a UKRI Future Leaders Fellow (MR/S03546X/1) and acknowledges funding from the UKRI Medical Research Council for the project Piloting A Secure Scalable Infrastructure for AI in the NHS (PASSIAN: MR/X005674/1).

The authors acknowledge members of the UCL POND group (http://pond.cs.ucl.ac.uk) for valuable feedback and input received during group discussions — in particular, Arman Eshaghi for suggesting the future work of using deep learning and robust registration.

We are grateful to the patients at the Essex Memory Clinic for agreeing to participate in research and to the Imaging Systems team of the Radiology Department at the Princess Alexandra Hospital for facilitating data access.

## References

[1] J. Garre-Olmo, “[Epidemiology of Alzheimer’s disease and other dementias],” (in spa), Rev Neurol, vol. 66, no. 11, pp. 377–386, Jun 01 2018. [Online]. Available: https://www.ncbi.nlm.nih.gov/pubmed/29790571.

[2] C. R. Jack et al., “NIA-AA Research Framework: Toward a biological definition of Alzheimer’s disease,” (in eng), Alzheimers Dement, vol. 14, no. 4, pp. 535–562, 04 2018, doi: 10.1016/j.jalz.2018.02.018.

[3] R. S. Desikan et al., “An automated labeling system for subdividing the human cerebral cortex on MRI scans into gyral based regions of interest,” (in eng), Neuroimage, vol. 31, no. 3, pp. 968–80, Jul 01 2006, doi: 10.1016/j.neuroimage.2006.01.021.

[4] B. Patenaude, S. M. Smith, D. N. Kennedy, and M. Jenkinson, “A Bayesian model of shape and appearance for subcortical brain segmentation,” (in eng), Neuroimage, vol. 56, no. 3, pp. 907–22, Jun 01 2011, doi: 10.1016/j.neuroimage.2011.02.046.

[5] L. Lenhart et al., “Anatomically Standardized Detection of MRI Atrophy Patterns in Early-Stage Alzheimer’s Disease,” (in eng), Brain Sci, vol. 11, no. 11, Nov 11 2021, doi: 10.3390/brainsci11111491.

[6] D. Archetti et al., “Inter-Cohort Validation of SuStaIn Model for Alzheimer’s Disease,” (in eng), Front Big Data, vol. 4, p. 661110, 2021, doi: 10.3389/fdata.2021.661110.

[7] K. Poulakis et al., “Heterogeneous patterns of brain atrophy in Alzheimer’s disease,” (in eng), Neurobiol Aging, vol. 65, pp. 98–108, 05 2018, doi: 10.1016/j.neurobiolaging.2018.01.009.

[8] P. Suppa et al., “Performance of Hippocampus Volumetry with FSL-FIRST for Prediction of Alzheimer’s Disease Dementia in at Risk Subjects with Amnestic Mild Cognitive Impairment,” (in eng), J Alzheimers Dis, vol. 51, no. 3, pp. 867–73, 2016, doi: 10.3233/JAD-150804.

[9] F. de Vos et al., “Combining multiple anatomical MRI measures improves Alzheimer’s disease classification,” (in eng), Hum Brain Mapp, vol. 37, no. 5, pp. 1920–9, May 2016, doi: 10.1002/hbm.23147.

[10] S. Alam, G.-R. Kwon, and T. A. s. D. N. Initiative, “Alzheimer disease classification using KPCA, LDA, and multi-kernel learning SVM,” International Journal of Imaging Systems and Technology, vol. 27, no. 2, pp. 133–143, 2017, doi: 10.1002/ima.22217.

[11] A. Bartos, D. Gregus, I. Ibrahim, and J. Tintěra, “Brain volumes and their ratios in Alzheimer’s disease on magnetic resonance imaging segmented using Freesurfer 6.0,” (in eng), Psychiatry Res Neuroimaging, vol. 287, pp. 70–74, 05 30 2019, doi: 10.1016/j.pscychresns.2019.01.014.

[12] J. D. Sluimer et al., “Accelerating regional atrophy rates in the progression from normal aging to Alzheimer’s disease,” (in eng), Eur Radiol, vol. 19, no. 12, pp. 2826–33, Dec 2009, doi: 10.1007/s00330-009-1512-5.

[13] E. Dicks et al., “Modeling grey matter atrophy as a function of time, aging or cognitive decline show different anatomical patterns in Alzheimer’s disease,” (in eng), Neuroimage Clin, vol. 22, p. 101786, 2019, doi: 10.1016/j.nicl.2019.101786.

[14] L. L. Backhausen, M. M. Herting, J. Buse, V. Roessner, M. N. Smolka, and N. C. Vetter, “Quality Control of Structural MRI Images Applied Using FreeSurfer-A Hands-On Workflow to Rate Motion Artifacts,” (in eng), Front Neurosci, vol. 10, p. 558, 2016, doi: 10.3389/fnins.2016.00558.

[15] T. G. Siddiqui et al., “Magnetic Resonance Imaging in Stable Mild Cognitive Impairment, Prodromal Alzheimer’s Disease, and Prodromal Dementia with Lewy Bodies,” (in eng), Dement Geriatr Cogn Disord, vol. 49, no. 6, pp. 583–588, 2020, doi: 10.1159/000510951.

[16] J. E. Iglesias et al., “Joint super-resolution and synthesis of 1 mm isotropic MP-RAGE volumes from clinical MRI exams with scans of different orientation, resolution and contrast,” (in eng), Neuroimage, vol. 237, p. 118206, 08 15 2021, doi: 10.1016/j.neuroimage.2021.118206.

[17] J. E. Iglesias et al., “SynthSR: A public AI tool to turn heterogeneous clinical brain scans into high-resolution T1-weighted images for 3D morphometry,” Science Advances, vol. 9, no. 5, p. eadd3607, 2023, doi: doi:10.1126/sciadv.add3607.

[18] M. Reuter, H. D. Rosas, and B. Fischl, “Highly accurate inverse consistent registration: a robust approach,” (in eng), Neuroimage, vol. 53, no. 4, pp. 1181–96, Dec 2010, doi: 10.1016/j.neuroimage.2010.07.020.

[19] D. L. Collins, P. Neelin, T. M. Peters, and A. C. Evans, “Automatic 3D intersubject registration of MR volumetric data in standardized Talairach space,” (in eng), J Comput Assist Tomogr, vol. 18, no. 2, pp. 192–205, 1994 Mar-Apr 1994.

[20] A. M. Dale, B. Fischl, and M. I. Sereno, “Cortical surface-based analysis. I. Segmentation and surface reconstruction,” (in eng), Neuroimage, vol. 9, no. 2, pp. 179–94, Feb 1999, doi: 10.1006/nimg.1998.0395.

[21] B. Fischl, M. I. Sereno, and A. M. Dale, “Cortical surface-based analysis. II: Inflation, flattening, and a surface-based coordinate system,” (in eng), Neuroimage, vol. 9, no. 2, pp. 195–207, Feb 1999, doi: 10.1006/nimg.1998.0396.

[22] B. Fischl, M. I. Sereno, R. B. Tootell, and A. M. Dale, “High-resolution intersubject averaging and a coordinate system for the cortical surface,” (in eng), Hum Brain Mapp, vol. 8, no. 4, pp. 272–84, 1999, doi: 10.1002/(sici)1097-0193(1999)8:4<272::aid-hbm10>3.0.co;2-4.

[23] B. Fischl and A. M. Dale, “Measuring the thickness of the human cerebral cortex from magnetic resonance images,” (in eng), Proc Natl Acad Sci U S A, vol. 97, no. 20, pp. 11050–5, Sep 26 2000, doi: 10.1073/pnas.200033797.

[24] B. Fischl, A. Liu, and A. M. Dale, “Automated manifold surgery: constructing geometrically accurate and topologically correct models of the human cerebral cortex,” (in eng), IEEE Trans Med Imaging, vol. 20, no. 1, pp. 70–80, Jan 2001, doi: 10.1109/42.906426.

[25] J. G. Sled, A. P. Zijdenbos, and A. C. Evans, “A nonparametric method for automatic correction of intensity nonuniformity in MRI data,” (in eng), IEEE Trans Med Imaging, vol. 17, no. 1, pp. 87–97, Feb 1998, doi: 10.1109/42.668698.

[26] B. Fischl et al., “Whole brain segmentation: automated labeling of neuroanatomical structures in the human brain,” (in eng), Neuron, vol. 33, no. 3, pp. 341–55, Jan 31 2002, doi: 10.1016/s0896-6273(02)00569-x.

[27] B. Fischl et al., “Automatically parcellating the human cerebral cortex,” (in eng), Cereb Cortex, vol. 14, no. 1, pp. 11–22, Jan 2004, doi: 10.1093/cercor/bhg087.

[28] A. Klein and J. Tourville, “101 labeled brain images and a consistent human cortical labeling protocol,” (in eng), Front Neurosci, vol. 6, p. 171, 2012, doi: 10.3389/fnins.2012.00171.

[29] M. Tanveer et al., “Machine Learning Techniques for the Diagnosis of Alzheimer’s Disease: A Review,” ACM Trans. Multimedia Comput. Commun. Appl., vol. 16, no. 1s, p. Article 30, 2020, doi: 10.1145/3344998.

[30] R. V. Marinescu, A. Eshaghi, D. C. Alexander, and P. Golland, “BrainPainter: A software for the visualisation of brain structures, biomarkers and associated pathological processes,” (in eng), Multimodal Brain Image Anal Math Found Comput Anat (2019), vol. 11846, pp. 112–120, Oct 2019, doi: 10.1007/978-3-030-33226-6_13.

[31] H. Braak, I. Alafuzoff, T. Arzberger, H. Kretzschmar, and K. Del Tredici, “Staging of Alzheimer disease-associated neurofibrillary pathology using paraffin sections and immunocytochemistry,” (in eng), Acta Neuropathol, vol. 112, no. 4, pp. 389–404, Oct 2006, doi: 10.1007/s00401-006-0127-z.

[32] C. J. Weber et al., “The Worldwide Alzheimer’s Disease Neuroimaging Initiative: ADNI-3 updates and global perspectives,” Alzheimer’s & Dementia: Translational Research & Clinical Interventions, vol. 7, no. 1, p. e12226, 2021, doi: 10.1002/trc2.12226.

[33] R. Raman et al., “Tackling a Major Deficiency of Diversity in Alzheimer’s Disease Therapeutic Trials: An CTAD Task Force Report,” The Journal of Prevention of Alzheimer’s Disease, vol. 9, no. 3, pp. 388–392, 2022/07/01 2022, doi: 10.14283/jpad.2022.50.

[34] N. P. Oxtoby, D. C. Alexander, and f. t. E. consortium, “Imaging plus X: multimodal models of neurodegenerative disease,” Current Opinion in Neurology, vol. 30, no. 4, pp. 371–379, 2017, doi: 10.1097/wco.0000000000000460.

[35] N. P. Oxtoby, “Data-Driven Disease Progression Modelling,” ed. arXiv:2211.05786, 2022.

[36] D. Ravi et al., “Degenerative adversarial neuroimage nets for brain scan simulations: Application in ageing and dementia,” (in eng), Med Image Anal, vol. 75, p. 102257, 01 2022, doi: 10.1016/j.media.2021.102257.

